# Race, Hypertensive Disorders of Pregnancy and Outcomes in Peripartum Cardiomyopathy

**DOI:** 10.1101/2023.10.29.23297730

**Authors:** Vincenzo B. Polsinelli, Agnes Koczo, Amber E. Johnson, Uri Elkayam, Leslie Cooper, John Gorcsan, Joan Briller, James Fett, Dennis M. McNamara, the IPAC investigators

## Abstract

**Introduction:** Black women with peripartum cardiomyopathy (PPCM) have a high prevalence of hypertensive disorders of pregnancy (HDP) and worse clinical outcomes compared with other races. We examined the role of HDP and race on myocardial recovery in women with PPCM.

**Methods:** A total of 100 women were enrolled into the Investigation in Pregnancy Associated Cardiomyopathy (IPAC) study. A hypertensive disorder of pregnancy (HDP) was defined as a gestational hypertension and preeclampsia, chronic hypertension was not included in the HDP subgroup. Left ventricular ejection fraction (LVEF) was assessed by echocardiography at entry, 6, and 12-months post-partum (PP). Clinical outcomes including persistent cardiomyopathy (LVEF≤35%), left ventricular assist device, (LVAD), transplantation, or death at 12-months were examined with and without HDP and between Black and non-Black subsets of women.

**Results:** Mean age in IPAC was 30±6 years, with a baseline LVEF of 35±10%. In Black women, those with HDP were more likely to present earlier (days PP HDP: 35±20 vs 54±27 days, P=0.03) compared to women without HDP. There was no difference in LVEF at study entry, but better recovery with HDP at 6 (HDP:0.52±0.11 vs no HDP:0.40±0.14, P=0.03) and 12-months (HDP:0.53±0.10 vs no HDP:0.40±0.16, P=0.02). At 12-months, Black women had a lower LVEF than non-Black women (P=0.007), driven by worse recovery in Black women without HDP (P=0.002). Black women with HDP had a similar LVEF to non-Black women (P=0.31).

**Conclusions:** In women with PPCM, HDP among Black women was associated with earlier presentation and better recovery compared to Black women without HDP.

## 1. Introduction

Peripartum cardiomyopathy (PPCM) is a rare complication of pregnancy, which remains a significant cause of morbidity and mortality. With an incidence in the United States of approximately 1 in 2000 live births, it is even greater cause of maternal mortality worldwide. (1,2) Hypertensive disorders of pregnancy (HDP) are a major risk factor for PPCM. Furthermore, HDP occur more frequently in Black women who incur an independent increased risk of PPCM. (2-4) The exact pathophysiology of PPCM remains uncertain, but several hypotheses exist such as immune, inflammatory, vascular, or genetic causes. (5-11) Recovery after PPCM, defined as a return of left ventricular ejection fraction (LVEF) >50%, is frequent and has been reported as up to 89% of women who had received optimized heart failure therapy. However 7-13% of patients are reported to experience adverse cardiac events including left ventricular assist device (LVAD), cardiac transplantation, or death. (12-14)

An accumulating volume of evidence suggests PPCM and HDP have a shared pathophysiology, and that PPCM in women with co-morbid HDP is driven by a shared mechanism. (ref), It is therefore reasonable to expect that these women may have similar outcomes and disease severity. Racial differences in PPCM have been described, although there is no consensus explaining differences observed.(15,16) Studies have demonstrated that Black women have worse outcomes compared to white counterparts.(16,17) These differences have been attributed to race-based health disparities including barriers to accessing care, delays in presentation, and higher rates of comorbid conditions such as diabetes, and greater risk of HDP.(18) Some cultural practices such as high salt ingestion in the perinatal period practiced by some regions of Nigeria where rates of PPCM are estimated around 1 in 100 live births have also been proposed, but failed to show statistical association.(19) In this study, we evaluated the role of race and HDP on disease severity, myocardial recovery, and outcomes for women with PPCM in the Investigations of Pregnancy Associated Cardiomyopathy (IPAC) study.

## 2. Methods

### 2.1 Study population

The IPAC multi-center registry enrolled 100 women with newly diagnosed PPCM within 13 weeks PP. (14) Enrollment criteria included 18 years of age or older, no history of cardiac disease, left ventricular ejection fraction (LVEF) ≤ 45% at the time of enrollment, and evaluation consistent with recent onset non-ischemic cardiomyopathy. Women with significant valvular disease, coronary disease, bacterial septicemia, ongoing drug or alcohol use disorders, history of chemotherapy, chest radiation within 5 years, or a history of previous cardiomyopathy were excluded.

### 2.2 Data collection

At the time of enrollment, demographic information including self-described race, pertinent delivery details, previous clinical evaluation, and current medical therapy were recorded. Medical therapy was recorded as drug class, dose was not recorded. Women were followed until 1 year PP. All hospitalizations and major cardiac events including implantation of an left ventricular assist device (LVAD), cardiac transplantation, or death were recorded. The study protocol was reviewed by the institutional review board of all participating centers, and informed consent was obtained from all subjects.

### 2.3 Echocardiographic assessment

Women enrolled in IPAC had an echocardiogram to assess LVEF at study entry, 6, and 12 months. Echocardiographic measurements including LVEF were assessed in a core laboratory (University of Pittsburgh). LVEF was calculated by biplane Simpson’s rule using a manual tracing of digital images. LV end-systolic and LV end-diastolic diameters (LVEDD) were assessed in the parasternal long-axis view and left atrial diameter (LAD) was assessed in the apical 4-chamber view.

### 2.4 Cohorts and subgroups

HDP were defined as having a diagnosis during the index pregnancy of pre-eclampsia, or gestational hypertension prior to the diagnosis of PPCM. Women with chronic hypertension (occurring prior to index pregnancy) were not included into the HDP subgroup, but were included in the no-HDP subgroup. Women who self-identified as Black or African American were included in the Black subgroup, and were compared with all other groups combined (women identifying as White, Asian or other).

### 2.5 Statistical Analysis

Differences in continuous variables between Black and HDP subgroups were assessed using ANOVA and two-way ANOVA. Bonferroni post-hoc testing was used to compare differences between subgroups. Differences in categorical variables between subgroups were calculated using chi-square. The Kaplan-Meier method was used to estimate time free from events, and the Log-Rank test was used to estimate differences between HDP and non-HDP subgroups. P-values <0.05 were considered statistically significant. Statistical analyses and graphics were completed using R, and Stata 12 (*Stata Statistical Software: Release 12*. College Station, TX: StataCorp LP.).

### 2.6 Outcomes

Persistent cardiomyopathy was defined as an LVEF ≤ 35% at 12 months post-partum, or last recorded LVEF. Survival free from major cardiovascular events (LVAD, transplantation, or death) was determined up to 12 months PP. Partial or full recovery was defined as LVEF>35%. A composite outcome of persistent cardiomyopathy, LVAD, transplantation, or death was used at 12 months.

## 3. Results

### 3.1 Baseline Characteristics

Mean age in the overall PPCM cohort was 30 ± 6 years, with a baseline LVEF of 34% ± 10. Differences between HDP subgroups are outlined in **Table 1**. Among non-Black women, women with HDP delivered via caesarian more often than did non-Black women without HDP (non-Black: HDP: 15/21 vs non-HDP: 22/49; P=0.04). Among Black women with HDP, there were significantly higher rates of breastfeeding and twin-gestation compared with Black women without HDP (Breastfeeding: Black: HDP=4/16 vs non-HDP=0/14; P=0.04, Twin gestation: Black: HDP=4/16 vs non-HDP=0/14 ; P=0.04). Medication prescription including guideline directed medical therapy for heart failure was similar between the four subgroups.

**Table 1.**
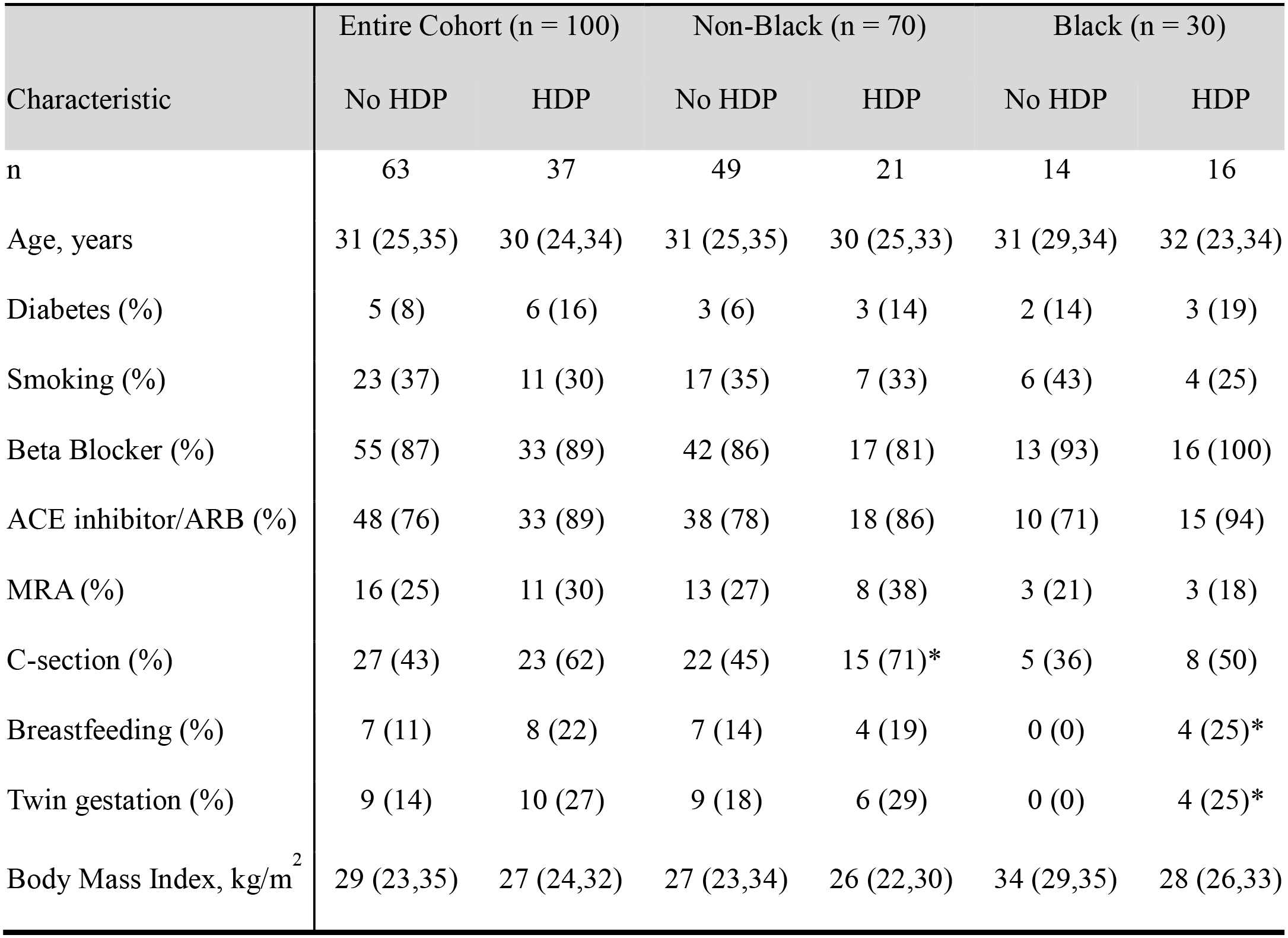
Clinical and demographic characteristics by HDP and race. Categorical variables are displayed as number (%), continuous variables as median and interquartile range (IQR), (*) denotes P-value <0.05. ACE = angiotensin converting enzyme, ARB = angiotensin receptor blocker, MRA = mineralocorticoid receptor antagonist.

| Characteristic | Entire Cohort (n = 100) |  | Non-Black (n = 70) |  | Black (n = 30) |  |
| --- | --- | --- | --- | --- | --- | --- |
|  | No HDP | HDP | No HDP | HDP | No HDP | HDP |
| n | 63 | 37 | 49 | 21 | 14 | 16 |
| Age, years | 31 (25,35) | 30 (24,34) | 31 (25,35) | 30 (25,33) | 31 (29,34) | 32 (23,34) |
| Diabetes (%) | 5 (8) | 6 (16) | 3 (6) | 3 (14) | 2 (14) | 3 (19) |
| Smoking (%) | 23 (37) | 11 (30) | 17 (35) | 7 (33) | 6 (43) | 4 (25) |
| Beta Blocker (%) | 55 (87) | 33 (89) | 42 (86) | 17 (81) | 13 (93) | 16 (100) |
| ACE inhibitor/ARB (%) | 48 (76) | 33 (89) | 38 (78) | 18 (86) | 10 (71) | 15 (94) |
| MRA (%) | 16 (25) | 11 (30) | 13 (27) | 8 (38) | 3 (21) | 3 (18) |
| C-section (%) | 27 (43) | 23 (62) | 22 (45) | 15 (71)* | 5 (36) | 8 (50) |
| Breastfeeding (%) | 7 (11) | 8 (22) | 7 (14) | 4 (19) | 0 (0) | 4 (25)* |
| Twin gestation (%) | 9 (14) | 10 (27) | 9 (18) | 6 (29) | 0 (0) | 4 (25)* |
| Body Mass Index, kg/m<sup>2</sup> | 29 (23,35) | 27 (24,32) | 27 (23,34) | 26 (22,30) | 34 (29,35) | 28 (26,33) |

### 3.2 Time to presentation

Differences in time to presentation post-partum are shown in **Figure 1**. Black women presented later than non-Black women (Black=43.0±25 days vs non-Black=26.6±23 days; P=0.0008), and women without HDP presented later than women with HDP (HDP=28.0±21 days vs no-HDP=33.5±26 days; P=0.02). Among women without HDP, Black women presented later than non-Black women (no-HDP: Black=53.6±27 days vs non-Black=27.8±24 days; P=0.02). However, among women with HDP, there was no difference between Black, and non-Black women (HDP: Black=33.6±21 days vs non-Black=23.8±21 days; P=1.00).

**Figure 1:**
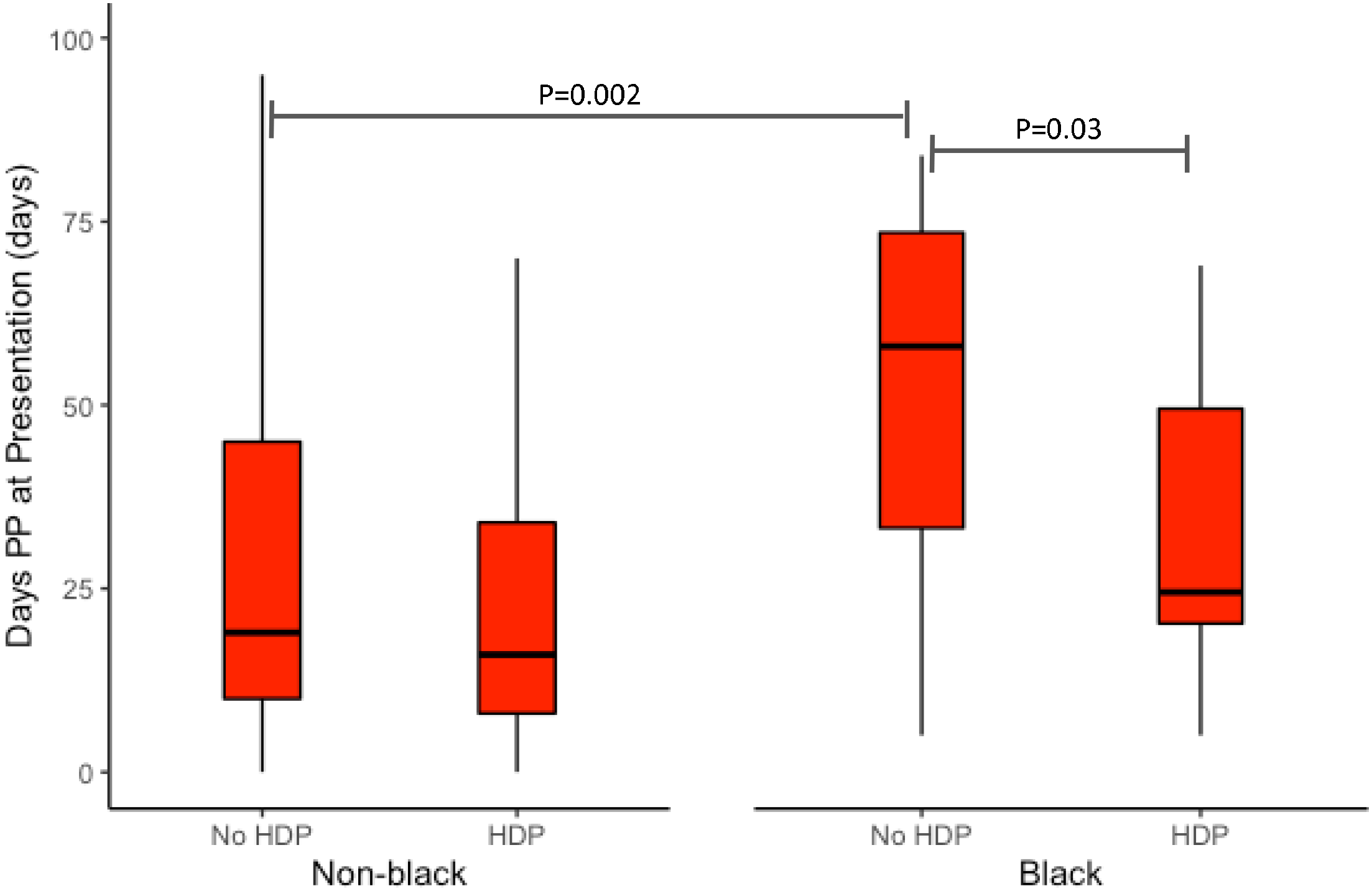
Days PP at study entry by race and HDP -- Boxplots represent the interquartile range from the first to the third quartile. Median represented by a thick Black line across the interquartile range, and error bars represent 1.5 times the interquartile range. Days PP are shown between Black and non-Black women and by subgroups based on a history of HDP during index pregnancy. Significant differences between pairs of subgroups are shown using vertical hash (|) to represent comparison group, connected by a horizontal bar. HDP = hypertensive disorder of pregnancy; PP = post-partum

### 3.3 Remodeling, and myocardial recovery

Assessment of myocardial recovery, by measurement of LVEF at enrollment, 6, and 12 months in each subgroup is shown in **Figure 2**.

**Figure 2:**
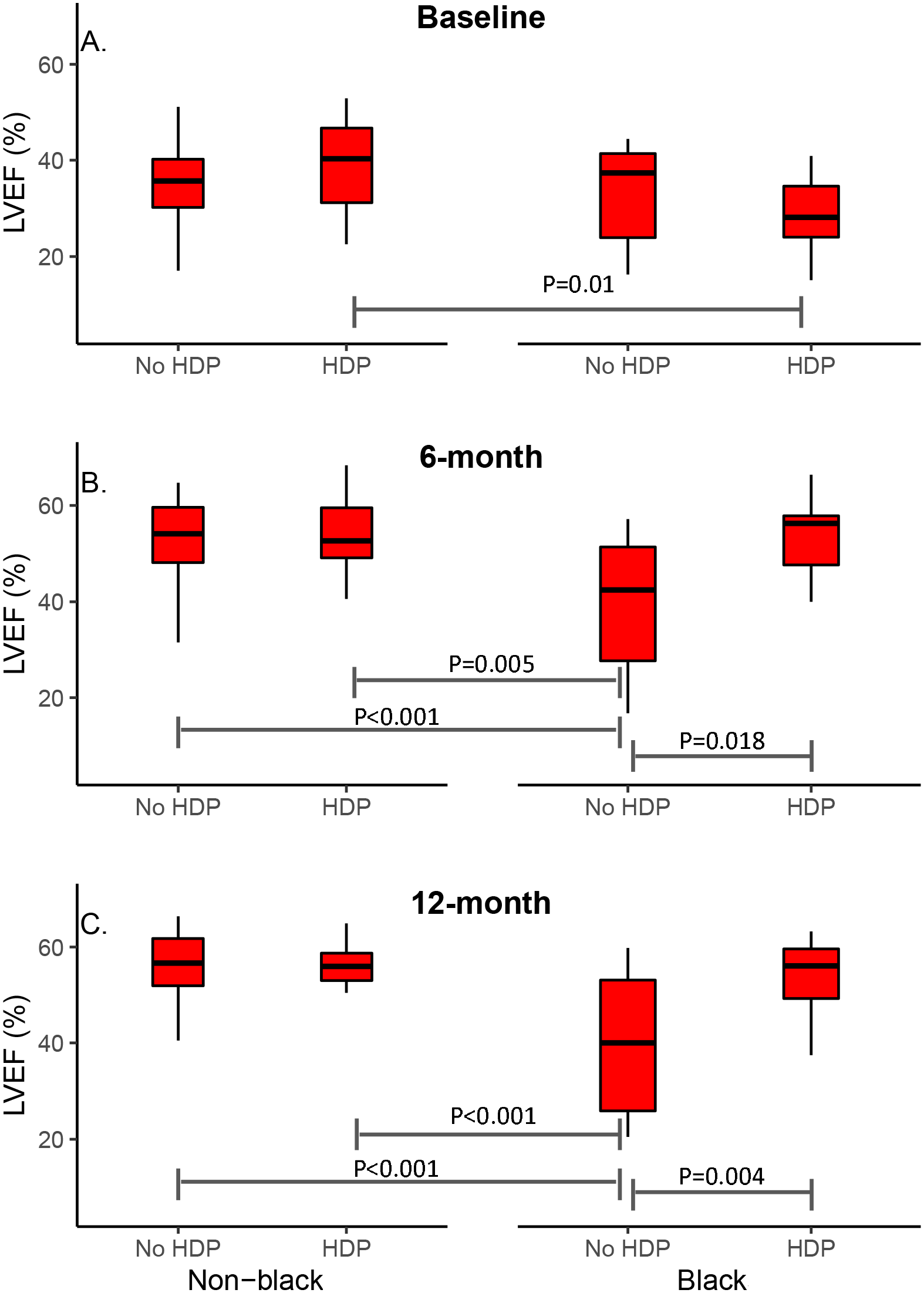
LVEF (%) by race and HDP at baseline, 6, and 12 months PP -- Boxplots represent the interquartile range from the first to the third quartile. Median represented by a thick Black line across the interquartile range, and error bars represent 1.5 times the interquartile range. LVEF is shown between Black and non-Black women and by subgroups based on a history of HDP during index pregnancy. (A) Baseline. (B) Six-months (C) Twelve-months. Significant differences between pairs of subgroups are shown using vertical hash (|) to represent comparison group, connected by a horizontal bar. LVEF = left ventricular ejection fraction; HDP = hypertensive disorder of pregnancy; PP = post-partum

#### Enrollment

At study enrollment, Black women had lower LVEF than non-Black (Black=30.6±9.2% vs non-Black=36.0±9.3%; P=0.005) but there was no difference between HDP and non-HDP subgroups (HDP=34.3±9.8% vs no-HDP=34.4±9.5%; P=0.91). Among women without HDP, there was no difference in LVEF at enrollment between women who were Black compared to non-Black (no-HDP: Black=32.8±10.4% vs non-Black=34.9±9.3%; P=0.43). However, among women with HDP, women who were Black had lower LVEF at time of enrollment (HDP: Black=28.7±7.8% vs non-Black=38.5±9.0%; P=0.01).

#### 6-month follow-up

At 6 months follow-up, Black women (Black=46.2±14.0% vs non-Black=53.3±8.2%; P=0.005) and women without HDP (no-HDP=50.5±10.9% vs HDP=52.6±10.3%; P=0.019) had lower LVEF. Among women without HDP, Black women had lower LVEF at 6 months compared to non-Black women (no-HDP: Black=39.6±14.1% vs non-Black=53.5±7.8%; P=0.005), however there was no race-based difference among women with HDP (HDP: Black=52.2±11.5% vs non-Black=53.0±9.6%; P=1.0).

#### 12-month follow-up

These differences persisted through the 1-year follow-up period by race (Black=46.9±14.3% vs non-Black=55.6±7.0%; P=0.0002) and HDP status (HDP=54.2±7.9% vs no-HDP=52.4±11.7%; P=0.009). Among women without HDP, Black women had lower LVEF at 12 months compared to non-Black women (no-HDP: Black=39.5±15.9% vs non-Black=55.9±7.2%; P<0.001), but there was no race-based difference among women with HDP(HDP: Black=53.1±9.7% vs non-Black=54.9±6.6%; P=0.55). Compared to all other groups, Black women without HDP had the worst rates of myocardial recovery at 12 months **(Figure 2)**.

#### Remodeling

Differences in cardiac remodeling including LVEDD and LAD in each subgroup are shown in **Figure 3**. Among patients who did not have HDP, patients who were Black had worse LVEDD compared to non-Black (no-HDP: Black=60±7mm vs non-Black=54±6mm; P=0.029). Among patients who were Black, patients without HDP had worse LAD compared to patients who did not have HDP (Black: HDP=37±7mm vs no-HDP=42±6mm; P=0.033). Overall, women who were Black and without HDP had worse LVEDD and LAD.

**Figure 3:**
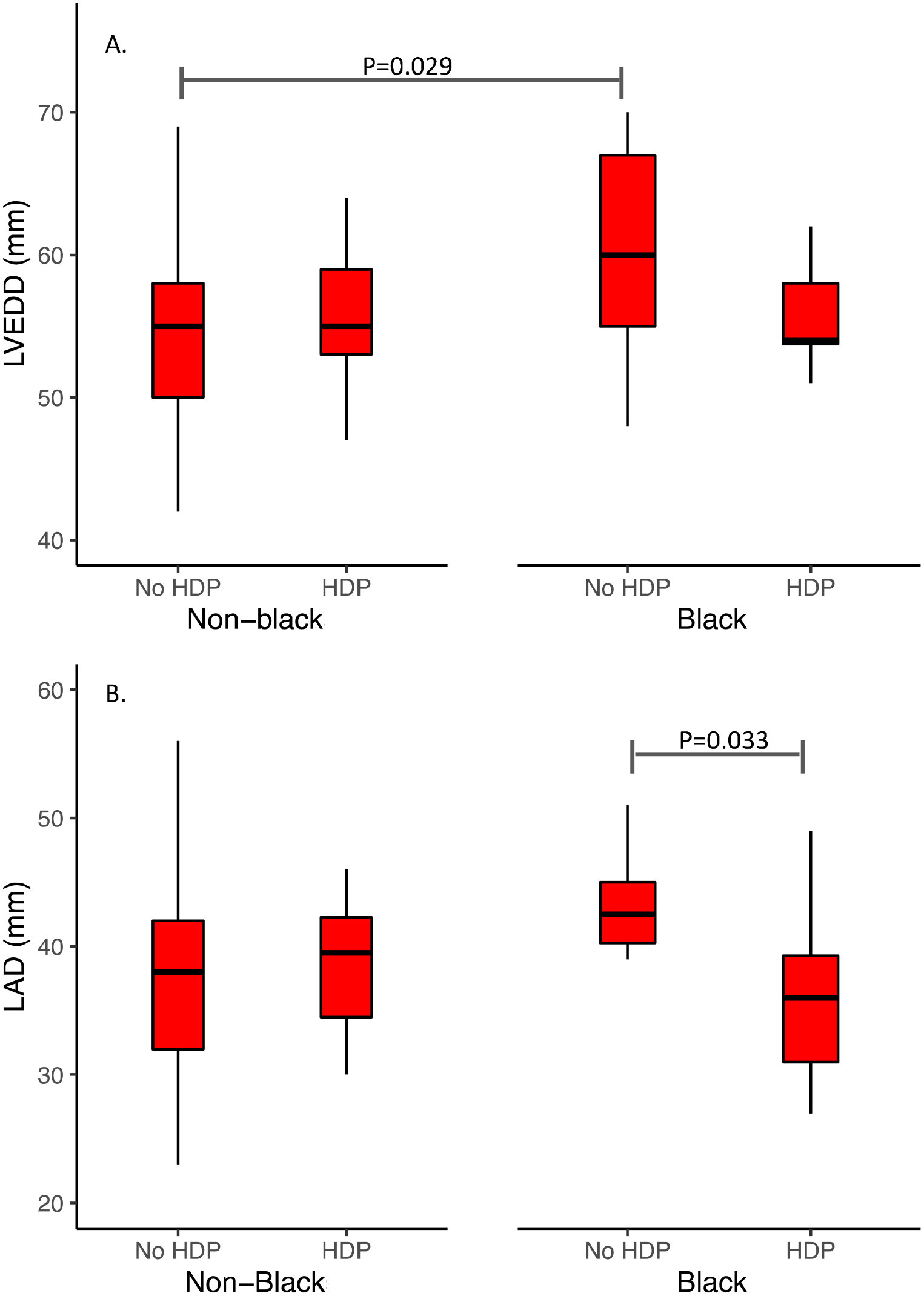
LVEDD (A) and LAD (B) (mm) by race and HDP at baseline -- Boxplots represent the interquartile range from the first to the third quartile. Median represented by a thick Black line across the interquartile range, and error bars represent 1.5 times the interquartile range. LVEDD and LAD are shown between Black and non-Black women and by subgroups based on a history of HDP during index pregnancy. Significant differences between pairs of subgroups are shown using vertical hash (|) to represent comparison group, connected by a horizontal bar. LVEDD = left ventricular end-diastolic diameter; LAD = left atrial diameter; HDP = hypertensive disorder of pregnancy.

### 3.4 Survival and outcomes

At the end of 12 months post-partum, 6 women had either required an LVAD or cardiac transplantation or died. Another 6 women had no recovery of LV function (LVEF ≤ 35% at 12 months). The number of women who met the composite end-point of persistent cardiomyopathy, LVAD, cardiac transplantation, or death, at the end of the 12 month follow-up period are shown in **Figure 4**. More Black women met the composite end-point than non-Black women (P=0.0008), and more women without HDP met the composite outcome than women with HDP (P=0.001) (**Figure 4)**. There was no difference in event-free survival in women with versus without HDP, however no women with HDP experienced an event of LVAD or cardiac transplantation, or death (Log-Rank; P=0.06).

**Figure 4.**
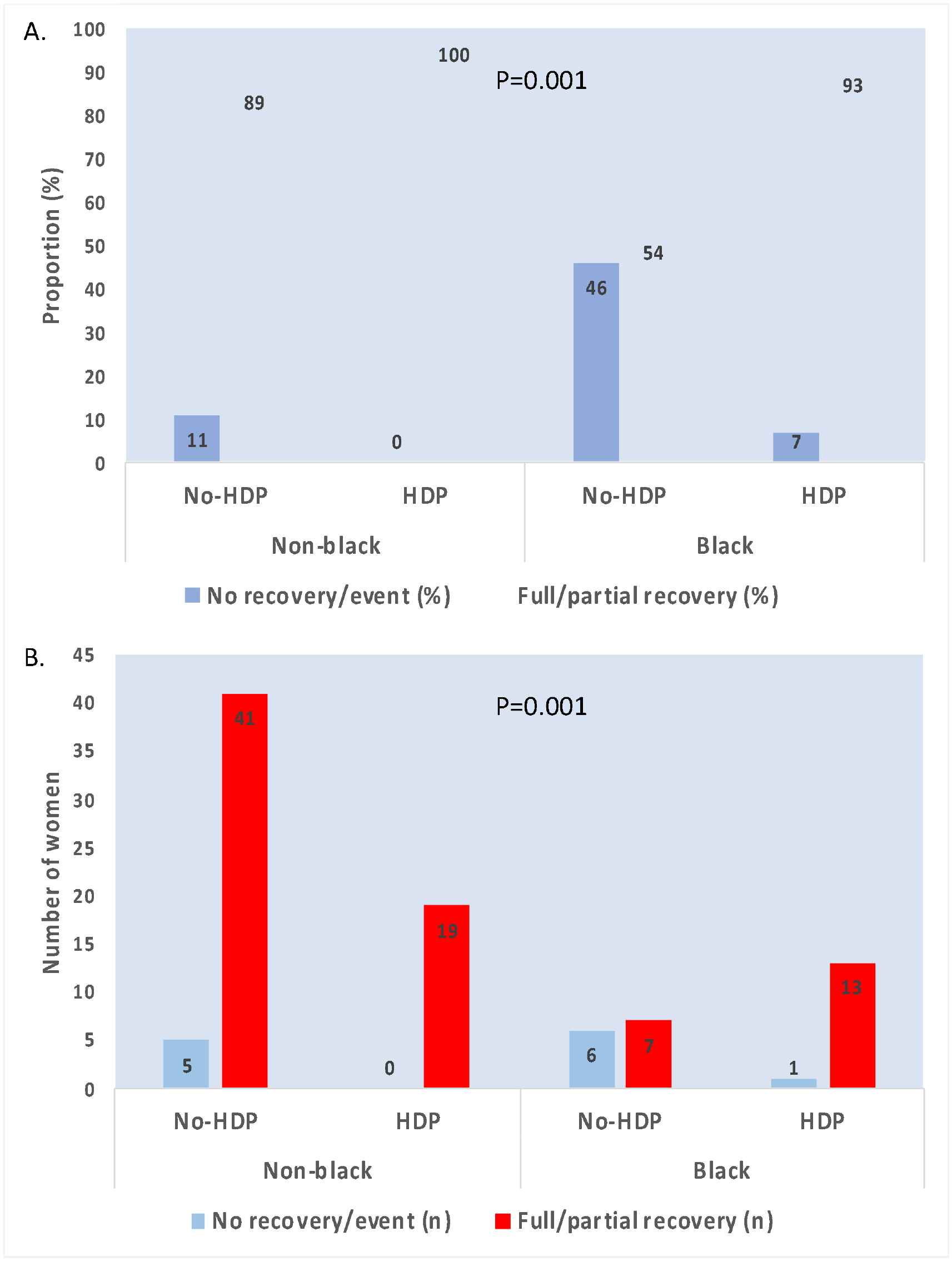
Number of women with full or partial recovery compared to, LVAD, transplant, death or persistent cardiomyopathy at 12 months PP by Race and HDP– Panel A bar graph represents the proportion of women in each subgroup at 12-months, and panel B represents the number of women in each subgroup at 12-months of follow up. The outcomes at 12-months are illustrated using colored bars, no recovery/event is defined as: persistent cardiomyopathy (LVEF ≤ 35%), LVAD, cardiac transplantation, or death. Full/partial recovery is defined as those with LVEF those who experienced a full or partial recovery (LVEF > 35%) at 12 months or last follow-up. Chi-squared between groups was used to calculate statistical significance. Post-hoc testing of adjusted residuals showed the subgroup non-HDP/Black was significantly different than other subgroups (P=0.00013). Statistical significance of all groups is shown in figure (P=0.001). LVAD = left ventricular assist device; PP = post-partum; LVEF = left ventricular ejection fraction; HDP = hypertensive disorder of pregnancy.

## 4. Discussion

In this study from IPAC, Black women without HDP had later presentation, more cardiac remodeling and lower rates of LVEF recovery. Moreover, these women had an increased risk of LVAD, cardiac transplantation, death, or persistent cardiomyopathy at 12 months compared to Black women with HDP, and non-Black women. In contrast, women with HDP had no difference in LVEF at time of enrollment, which persisted through follow-up at 12 months as compared to their non-HDP counterparts. In addition, women with HDP were significantly less likely to experience a composite event of, LVAD, cardiac transplantation death or persistent cardiomyopathy at the conclusion of the 12 month follow up period. Prior studies have suggested both race and HDP have impacts on LVEF recovery and outcomes in PPCM. (15,20) Our study suggests HDP may attenuate racial differences seen in PPCM, possibly by facilitating an earlier diagnosis which may improve myocardial recovery and outcomes in this population.

### 4.1 Myocardial recovery and outcomes

These data are concordant with several studies which demonstrate worse myocardial recovery and outcomes among Black women with PPCM. (16,20,21) Observations of racial differences in this cohort have been discussed previously.(14) Reports from Germany and Japan from mostly non-Black cohorts have observed better recovery and LVEF among women with HDP. (22,23) Our study found the presence of HDP among Black women was associated with earlier presentation and better outcomes compared to Black women without HDP. Similar trends were observed from two studies of Black women from South Africa which showed an association of higher baseline systolic blood pressure in survivors compared to non-survivors. However, these studies excluded women with hypertension, and speculated lower baseline blood pressure may be related to cardiogenic shock rather than an absence of HDP. (24,25)

There are several possible explanations for our findings. The earlier presentation of patients with HDP may be explained by the observation that developing HDP during pregnancy may facilitate overall closer follow up care including into the postpartum setting, thus potentially attenuating any barriers to accessing medical care. Lewey et. al. showed delayed time to diagnosis was associated with worse outcomes among women with PPCM, a finding that was maintained in Black and non-Black subgroups irrespective of HDP status. (20) It is well known that earlier initiation of evidence-based therapies for heart failure is associated with improved cardiac remodeling and longer survival. Although prompt diagnosis is necessary before intentional prescription of evidence-based therapies such as beta-blockers or angiotensin-converting enzyme inhibitors, pharmacologic management of HDP in the peripartum period includes medicines from the same drug classes. It is possible that women with HDP may have been initiated on the same or similar medicines that would be used for guideline-directed medical therapy in heart failure, for management of HDP. Our study found no difference in heart failure medications, however patients’ medications prior to diagnosis and titration during the study period were not assessed.

### 4.2 Strengths and Limitations

There are several strengths and limitations that must be discussed. A dedicated echo core lab which was used to adjudicate all echo measurements for the study. In addition, this study represents the largest US registry of patients with PPCM, of which risk factors and general demographics were well representative of the general US population (30% Black individuals, 37% with HDP). The IPAC study was conducted across 30 different centers throughout the United States. This imposes heterogeneity in practice patterns and patient management which was left to the discretion of the managing physician. Detailed information regarding medical therapy was not available including doses of HF therapies. Further, although this is the largest US registry of patients with PPCM, it is still a small sample size and lacks statistical power.

## 4.2 Conclusions

This study from IPAC demonstrated a relationship between Black women, and HDP status on myocardial recovery and outcomes in patients with PPCM. We demonstrated Black women without HDP have later presentation, lower LVEF at baseline, were less likely to have myocardial recovery. Furthermore, this subset of women had worse remodeling including greater LAD, and greater LVEDD. Black women without HDP were also observed to have higher clinical composite outcomes (, LVAD, cardiac transplantation death or persistent cardiomyopathy at 12 months) compared to all other subgroups. However Black women with HDP showed no difference in time to presentation, 12-month LVEF, or composite outcome when compared to women who were non-Black. Together, these data suggest the presence of HDP may attenuate the racial differences observed in PPCM. Further studies should investigate possible mechanisms of this finding. Our findings also suggest that early diagnosis and treatment for women at risk PPCM could help prevent adverse outcomes and promote more equitable care for racially diverse patient populations.

## Data Availability

All data produced in the present study are available upon reasonable request to the authors

## Abbreviations

(PPCM): Peripartum cardiomyopathy
(HDP): Hypertensive disorders of pregnancy
(LVEF): Left ventricular ejection fraction
(PP): Post-partum
(LVEDD): Left Ventricular End-Diastolic Diameter
(LAP): Left Atrial Pressure
(IPAC): Investigation in pregnancy associated cardiomyopathy.

## References

1. Arany Z, Elkayam U. Peripartum cardiomyopathy. Circulation 2016;133:1397–1409.

2. Gunderson EP, Croen LA, Chiang V, Yoshida CK, Walton D, Go AS. Epidemiology of peripartum cardiomyopathy: incidence, predictors, and outcomes. Obstetrics & Gynecology 2011;118:583–591.

3. Behrens I, Basit S, Lykke JA et al. Hypertensive disorders of pregnancy and peripartum cardiomyopathy: a nationwide cohort study. PloS one 2019;14:e0211857.

4. Bello N, Rendon ISH, Arany Z. The relationship between pre-eclampsia and peripartum cardiomyopathy: a systematic review and meta-analysis. Journal of the American College of Cardiology 2013;62:1715–1723.

5. Ansari AA, Fett JD, Carraway RE, Mayne AE, Onlamoon N, Sundstrom JB. Autoimmune mechanisms as the basis for human peripartum cardiomyopathy. Clin Rev Allergy Immunol 2002;23:301–24.

6. Bültmann BD, Klingel K, Näbauer M, Wallwiener D, Kandolf R. High prevalence of viral genomes and inflammation in peripartum cardiomyopathy. Am J Obstet Gynecol 2005;193:363–5.

7. Lamparter S, Pankuweit S, Maisch B. Clinical and immunologic characteristics in peripartum cardiomyopathy. Int J Cardiol 2007;118:14–20.

8. Sheppard R, Hsich E, Damp J et al. GNB3 C825T Polymorphism and Myocardial Recovery in Peripartum Cardiomyopathy: Results of the Multicenter Investigations of Pregnancy-Associated Cardiomyopathy Study. Circ Heart Fail 2016;9:e002683.

9. Ware JS, Li J, Mazaika E et al. Shared Genetic Predisposition in Peripartum and Dilated Cardiomyopathies. N Engl J Med 2016;374:233–41.

10. Damp J, Givertz MM, Semigran M et al. Relaxin-2 and Soluble Flt1 Levels in Peripartum Cardiomyopathy: Results of the Multicenter IPAC Study. JACC Heart Fail 2016;4:380–8.

11. Patten IS, Rana S, Shahul S et al. Cardiac angiogenic imbalance leads to peripartum cardiomyopathy. Nature 2012;485:333–8.

12. Irizarry OC, Levine LD, Lewey J et al. Comparison of Clinical Characteristics and Outcomes of Peripartum Cardiomyopathy Between African American and Non-African American Women. JAMA Cardiol 2017;2:1256–1260.

13. Goland S, Bitar F, Modi K et al. Evaluation of the clinical relevance of baseline left ventricular ejection fraction as a predictor of recovery or persistence of severe dysfunction in women in the United States with peripartum cardiomyopathy. J Card Fail 2011;17:426–30.

14. McNamara DM, Elkayam U, Alharethi R et al. Clinical outcomes for peripartum cardiomyopathy in North America: results of the IPAC Study (Investigations of Pregnancy-Associated Cardiomyopathy). Journal of the American College of Cardiology 2015;66:905–914.

15. Getz KD, Lewey J, Tam V et al. Neighborhood education status drives racial disparities in clinical outcomes in PPCM. American Heart Journal 2021;238:27–32.

16. Kao DP, Hsich E, Lindenfeld J. Characteristics, adverse events, and racial differences among delivering mothers with peripartum cardiomyopathy. JACC Heart Fail 2013;1:409–16.

17. Goland S, Modi K, Hatamizadeh P, Elkayam U. Differences in clinical profile of African-American women with peripartum cardiomyopathy in the United States. Journal of cardiac failure 2013;19:214–218.

18. Gambahaya ET, Minhas AS, Sharma G et al. Racial Differences in Delivery Outcomes among Women with Peripartum Cardiomyopathy. CJC Open 2021.

19. Karaye KM, Ishaq N, Sa’idu H et al. Incidence, clinical characteristics, and risk factors of peripartum cardiomyopathy in Nigeria: results from the PEACE Registry. ESC heart failure 2020;7:236–244.

20. Lewey J, Levine LD, Elovitz MA, Irizarry OC, Arany Z. Importance of Early Diagnosis in Peripartum Cardiomyopathy. Hypertension 2020;75:91–97.

21. Sinkey RG, Rajapreyar IN, Szychowski JM et al. Racial disparities in peripartum cardiomyopathy: eighteen years of observations. J Matern Fetal Neonatal Med 2020:1–8.

22. Haghikia A, Podewski E, Libhaber E et al. Phenotyping and outcome on contemporary management in a German cohort of patients with peripartum cardiomyopathy. Basic research in cardiology 2013;108:366.

23. Kamiya CA, Kitakaze M, Ishibashi-Ueda H et al. Different characteristics of peripartum cardiomyopathy between patients complicated with and without hypertensive disorders– results from the Japanese nationwide survey of peripartum cardiomyopathy–. Circulation Journal 2011:1105251244–1105251244.

24. Sliwa K, Förster O, Libhaber E et al. Peripartum cardiomyopathy: inflammatory markers as predictors of outcome in 100 prospectively studied patients. European heart journal 2006;27:441–446.

25. Blauwet LA, Libhaber E, Forster O et al. Predictors of outcome in 176 South African patients with peripartum cardiomyopathy. Heart 2013;99:308–313.

